# Effect of Complete Respiratory Muscle Training (cRMT) on Dysphonia following Single CVA: A retrospective pilot study

**DOI:** 10.1101/2020.12.20.20248589

**Authors:** Robert J. Arnold, Christopher S Gaskill, Nina Bausek

## Abstract

**Background:** Although dysphonia is less prevalent than dysphagia following cerebrovascular accidents, dysphonia does contribute to the burden of disease resulting from stroke. Strengthening muscles of the larynx and respiratory tract through respiratory muscle training (RMT) has proven effective in improving voice after neurological insult. However, approaches to strengthen only the expiratory muscle groups (EMST) dominate the clinical study literature, with variable outcomes. By focusing on exhalation, the contribution of inspiratory muscles to phonation may have been overlooked. This study investigated the effect of combined respiratory muscle training (cRMT) to improve voice function in stroke patients.

**Methods:** Recorded data of twenty patients with dysphonia following stroke were allocated to an intervention (IG) or a control group (CG) based upon whether they chose cRMT or not while awaiting *pro bono* voice therapy services. The intervention group (N=10) was treated daily with three 5-minute sessions of complete resistive respiratory muscle training for 28 days, while the control group (N=10) received no cRMT or other exercise intervention. Perceptual and acoustic measurements as well as a pulmonary function test were assessed pre-and post-intervention.

**Results:** The intervention group demonstrated significant improvements after 28 days of combined respiratory muscle training (cRMT) in peak flow (127%), patient self-perception of voice improvement (84.41%), as well as in five of the six categories of the Consensus Auditory-Perceptual Evaluation of Voice (CAPE-V) overall severity (63.22%), breathiness (61.06%), strain (63.43%), pitch range (48.11%) and loudness (57.51%), compared to the control group who did not receive treatment. Furthermore, cRMT also led to significant improvements in maximum phonation time (212.5%) and acoustic parameters of vocal intensity and semitone range.

**Conclusion:** This pilot study shows promise of the feasibility and effectiveness of cRMT to lessen the signs and symptoms of dysphonia while simultaneously improving breath support.

## Introduction

Voice disorders can have multiple underlying etiologies including structural, phonotraumatic, functional, neurogenic, and iatrogenic pathologies. Voice disorders are present when “quality, pitch, loudness, or flexibility differ from the voices of others of similar age, sex, and cultural group”[1]. In practice, a voice disorder exists when the perceptual characteristics of an individual’s voice create sufficient distraction to the extent the listener attends more to the sound of the individual’s voice rather than the content and emotion s/he is conveying. Dysphonia, also commonly referred to as an impairment of voice production, has been reported to affect nearly one third of all people at one point or another across their lifespan [2]. Neurogenic dysphonia resulting from stroke has been traditionally assessed and treated clinically as a feature of dysarthria (a motor speech disorder). However, for the purposes of this study, we consider neurogenic dysphonia resulting from a cerebrovascular accident (CVA) as a unique disorder.

The overall prevalence of voice disorders in the US is increasing, with an annual incidence of voice disorders of 7.6% [3,4]. Incidence and prevalence of voice disorders in the elderly are not well established, with an observed prevalence ranging from 2.5% to 6% in individuals over age 70, based on insurance claims data [5,6]. Prevalence in the institutionalized elderly may exceed those reports, with an observed prevalence of 20% of perceived dysphonia. The marked reduction in quality of life reported by 50% of elderly may be associated with the voice disorder directly, as well as with the frequently present comorbidities, primarily pulmonary disease and neurological illness [7,8].

Incidence and prevalence of dysphonia following a single CVA is more difficult to determine due to the diagnostic overlap with dysarthria, as well as the co-occurrence of dysarthria with dysphagia, which tends to receive more attention in acute and subacute treatment settings [9,10]. Only one study to our knowledge has addressed the occurrence of vocal cord paresis following acute ischemic stroke [11], reporting a 20.4% incidence of vocal paresis in 54 patients. Vocal fold paresis was seen in all patients with a brainstem lesion, in 11.7% of patients with lacunar infarcts, and in 16.4% of patients with cortical and subcortical infarcts. The vocal cord paresis was significantly correlated with the presence of dysphonia.

While phonation has been defined as “sound generation by means of vocal fold vibrations” [12], the sound of a given voice is then further modified via vocal tract filtering achieved by changing the shape of the oropharynx. Physiologically, the process of phonation is a complex set of neuromuscular events coordinated by the nervous system, involving muscles of the pulmonary system and larynx. Adequate respiratory muscle function and airflow are essential for voice production and phonation [13]. Both the muscles of inhalation as well as the muscles of exhalation are critical integrated components of the upper aerodigestive tract, whose function is necessary for vocal physiology [14]. Strengthening of either muscle group by inspiratory muscle strength training (IMST) or expiratory muscle strength training (EMST), respectively, may show positive effects on respiration as well as on vocal function [15–19]. Some studies have also shown an exercise regimen which includes EMST in the treatment plan to be beneficial in maximizing phonation in some neurological patient populations [20,21]. However, data on the effect of combined inspiratory and expiratory muscle strength training specifically on dysphonia has been lacking so far. Phonation is dependent upon a synergy between the action of both muscles of inspiration and expiration, to both provide adequate lung volume for voicing and to control the expiratory flow to meet the prosodic demands of connected speech. Therefore, it is important to understand this coordinated relationship and how voice can be affected through systematic training of both inspiratory and expiratory musculature. As the muscles of inspiration and expiration also play a critical role in supporting the upper aerodigestive tract’s swallow function, a recent study of interest examined the effects of a cRMT protocol on dysphagia after stroke and reported some positive effects on improving airway safety with swallowing [22].

To appreciate the potentially significant impact of CVA on voice, it is necessary to consider the many direct and indirect effects a CVA can have on phonation. A stroke can affect the phrenic nerve which controls the diaphragm as well as multiple cranial nerves that control speech and voice. Many additional indirect factors can contribute to dysphonia following CVA, including poor pulmonary reserve, general deconditioning, depression, polypharmacy, sequelae of intubation, poor secretion management, reduced laryngeal sensation, and gastroesophageal reflux [23]. Some pathophysiology literature indicates disuse of muscles results in generalized atrophy as well as a loss of muscle strength at a rate of around 12% a week [24]. Dysphonia has been shown to be a significant contributor to poor quality of life in this population, but often is either overlooked or undertreated due to the clinical treatment hierarchy of laryngeal functions which are, in order of priority, airway protection, respiration, and phonation.

The consequences of neurogenic dysphonia are varied. Vocal fold weakness and/or paralysis underlying dysphonia post stroke may cause glottic closure deficits that can result in impaired abilities of the muscles of the thoracic cage and secondary diminished capacity to bear down on the muscles of the abdomen. These subsequent physiological deficits in turn can result in impaired abilities with defecation, urination, parturition, cough, and throat clearing. Likewise, impaired glottic closure has been shown to result in diminished strength of the extremities [1]. Some other consequences correlated with neurogenic dysphonia include compromised mental health with depression and anxiety [25], occupational consequences, and personal as well as societal financial consequences [26].

The purpose of this study was to determine if select measures of vocal and pulmonary function significantly improved after an exercise program using a combined resistive IMST and EMST device. The device selected for the exercises was The Breather® (PN Medical Inc., US Patent Number 4,739,987)[27], which allows for simultaneous strengthening of the muscles of inhalation and exhalation [28,29]. This retrospective pilot study analyzes the effectiveness of a 4-week combined respiratory muscle training (cRMT) program, which was shown to be effective for the treatment of neurogenic dysphagia [22], on voice function in stroke patients with diagnosed dysphonia following a single cerebrovascular accident (CVA).

## Methods

This retrospective study analyzes data collected at a *pro bono* clinical speech-language pathology (SLP) clinic, operated by the Office of Hispanic Ministry at a Catholic Church in the Greater Birmingham Area of Alabama between 1998 and 2009. An Institutional Review Board (IRB) exemption for this study was granted by the Western Institutional Board prior to the review and analysis of the existing data. All participants included in the study presented with dysphonia following a single CVA. As there was a lag between diagnosis and available treatment opportunity, patients were offered a choice to be instructed in cRMT which included explanation, demonstration, and return demonstration. Patients who chose to participate in the cRMT protocol were included in the intervention group (IG), while those who chose not to use the device were considered as the control group (CG) for analysis.

Participants included in the analysis had no prior neurological history, no history of dysphonia prior to their CVA, no pharmacological or surgical interventions, and no other therapy for speech or voice during the study period. Furthermore, no other allied health therapy services which may have resulted in some degree of improvement of vocal fold functioning (e.g. occupational therapy, physical therapy, respiratory therapy, music therapy, etc.) were received during the study time period. Lastly, no changes in medications (e.g. changes in dosage, discontinuation, addition, etc.) were noted during the study time period. Exclusion criteria were failure to meet one or more of the inclusion criteria. The participants’ demographic data were reported by age, sex, race, hemisphere of CVA, and the presence of a single CVA without extension. A four-week home-based intervention of combined respiratory muscle training (cRMT) using the device was offered to all patients awaiting treatment.

### cRMT Intervention

Combined respiratory muscle training (cRMT) was administered using The Breather®. Patients were instructed to wear a nose clip, sit upright, and to forcefully inhale and exhale through the device using diaphragmatic breathing. Patients unable to maintain a tight lip seal with the mouthpiece of the cRMT device were trained to utilize a CPR (cardiopulmonary resuscitation) facemask in lieu of the standard mouthpiece. The duration of intervention was 28 days (4 weeks), and included three sessions per day. Each week, one of the sessions was supervised. Each session consisted of 3 sessions per day of up to 5 minutes of cRMT per session, as tolerated. Tolerance was judged by the patient’s report of fatigue or dizziness and the clinician’s subjective observations of patient fatigue. cRMT intensity was defined as the highest tolerated settings for both inhalation and exhalation. Importantly, these settings were set independently from each other. Compliance and training adherence were monitored via patient communication during weekly skilled intervention sessions. The control group received no treatment during the study duration.

### Assessments

To assess pulmonary function, we assessed peak flow using a peak flow meter (Spir-O-Flow Peak Flow Pocket Monitor, Spirometrics) by taking the average of 3 trials as there is a relationship between peak flow and phonation. Patient self-perception of their voice was assessed using a visual analogue scale (VAS) with a 100 mm horizontal line where “0” on the far left represented “I have no voice” and “100” on the far right represented “My voice is fine”. Participants judged their own voice both pre- and post-treatment period by placing a vertical hash mark somewhere along the horizontal line. The clinician then measured the distance in millimeters from the left end of the horizontal line to the hash mark, so the voice self-perception data is presented as a number from 0 to 100. Perceptual components of voice (overall severity, roughness, breathiness, strain, and loudness) were assessed using the Consensus Auditory-Perceptual Evaluation of Voice (CAPE-V), which employs a similar set of visual analog scales [30]. The component of pitch as included in the CAPE-V was omitted by the clinician due to the reason the CAPE-V instructions specify “scale rates whether the individual’s pitch deviates from normal for that personas gender, age, and referent culture” but excludes gender identification, hormonal treatment, or other reasons for what otherwise may be perceived as an abnormal speaking pitch. Instead, pitch range was added as an augmentation to the CAPE-V for the perceptual evaluation of voice due to the reason pitch variability in connected speech is integral for a speaker to convey the message of emotion simultaneously with the message of content [31–33]. Acoustic measurements were assessed using Lee Silverman Voice Therapy Companion (LSVTC) Software (LSVT Global, V.1.0.4, Boulder, CO) [34].

### Data Analysis

The primary research question of effectiveness of cRMT upon neurogenic dysphonia following stroke was examined with six multivariate 2 × 2 factorial analyses of variance with repeated measures. The between-subjects independent variable was group (control vs. intervention) and the within-subjects independent variable was time (pre-test vs. post-test). The first MANOVA had two dependent variables, average peak flow reading and the self-perception of voice using a VAS. The second MANOVA had six dependent variables, i.e., the six dimensions of the CAPE-V (overall severity, roughness, breathiness, strain, pitch range, and loudness). The third MANOVA had two dimensions of the average maximum duration of /a/ (db SPL and duration in seconds). The fourth MANOVA had three dimensions of the high /a/ task (db SPL, high frequency in Hz, and modal F_0_ (Hz)). The fifth MANOVA had three dimensions of the low /a/ task (db SPL, frequency in Hz, and modal F_0_ (Hz)). The sixth and final MANOVA had the three final dimensions of the LSVTC Clinician Software examining the vocal intensity at the functional phrase, text/word reading, and conversation levels). Significant multivariate analyses were followed by univariate analyses. Rejection of the null hypothesis that post-intervention outcomes did not differ between intervention and control group was assessed using post-hoc *t*-tests.

In order to accurately determine any meaningful changes in pitch range from pre- to post-test, the high and low frequency changes, as well as the changes in the total range from each subject were used to calculate changes in semitones. The conversion from a frequency range to a semitone range was calculated by entering the values in Hertz to the semitone converter found at http://www.homepages.ucl.ac.uk/~sslyjjt/speech/semitone.html [35].

## Results

### Demographic Data

Review of 116 patient records resulted in elimination of 96 subjects due to failure to meet the inclusion criteria. Data from twenty participants who met the inclusion criteria were included in this study. Twelve of these (60%) were females and eight males (40%). Among the ten participants in the control group, seven were female and three were male. Among the intervention group, five were female and five were male. The mean age for the participants was 60.7 years (*SD* = 11.53). The mean age of participants in the control group was 66.0 years (*SD* = 10.83) and the mean age for the participants in the intervention group was 55.3 years (*SD* = 9.98). The mean age of the intervention group was significantly less than the control group, *t* (18) = 2.30, *p* = .034.

### Assessment of peak flow and perception of voice

Adequate airflow is required for voice production and phonation. We therefore analyzed the impact of cRMT in peak expiratory flow, compared to no intervention in the control group. Four weeks of cRMT increased peak average flow by 127%, compared to 15.68% in the control group (Table 2, Figure 1a). The observed improvement in the intervention group was significantly higher than that in the control group (p < .01). Table 2 also illustrates that the patient self-perception of voice significantly changed over time in the intervention group by 84.41%, compared to a 4.85% increase in the control group (p < .05)(Figure 1b).

**Table 1:**
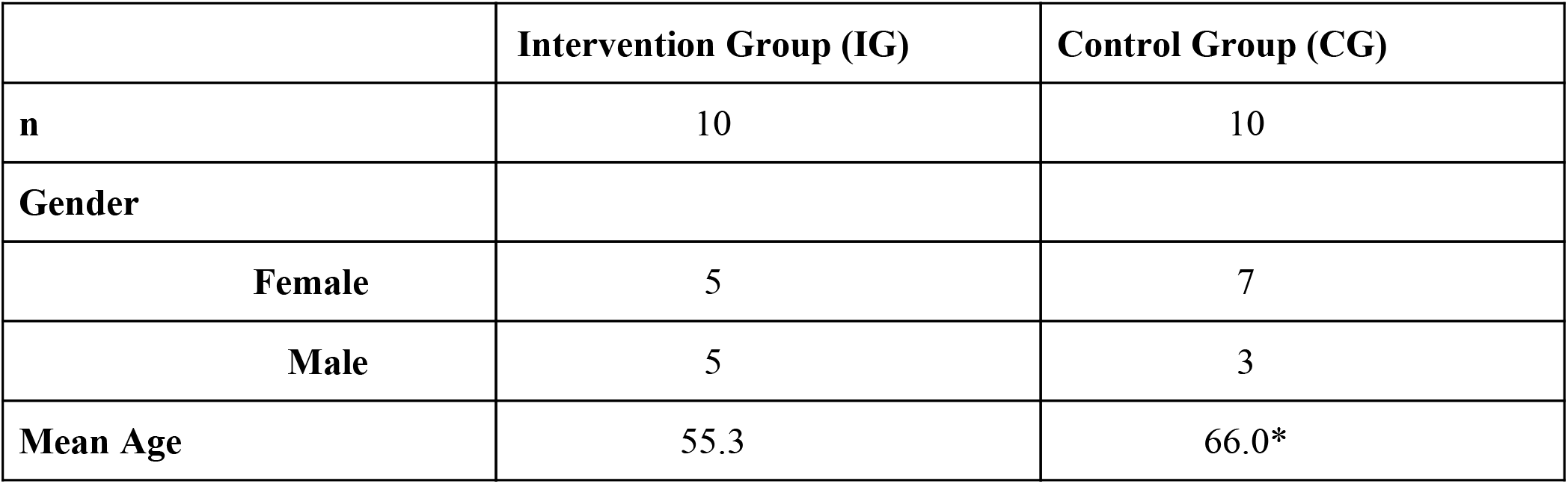
Patient demographics: Distribution of gender and mean age in the intervention group (IG) and control group (CG). *=Significant difference.

**Table 2:** Pre-test and post-test means and standard error (SEM), and percent change of peak flow and visual analog scale (VAS). Peak expiratory flow and visual analog scale of perception of voice were measured before (pre-test) and after (post-test) four weeks of cRMT (intervention group, IG) or no intervention (control group, CG). * indicates a significant post-test difference between intervention and control group. LPM = liters per minute, mm = millimeters.

|  | Group | Pretest |  | Post-test |  |  |  |
| --- | --- | --- | --- | --- | --- | --- | --- |
|  |  | Mean | SEM | Mean | SEM | Percent Change | p-Value |
| **Peak Flow (LPM)** | CG | 91.20 | 10.06 | 105.50 | 10.90 | 15.68% |  |
|  | IG | 92.60 | 11.05 | 210.20 | 25.81 | 127%* | <.01 |
| **VAS (mm)** | CG | 39.20 | 7.15 | 41.10 | 7.50 | 4.85% |  |
|  | IG | 37.20 | 6.75 | 68.60 | 8.89 | 84.41%* | <.05 |

**Figure 1:**
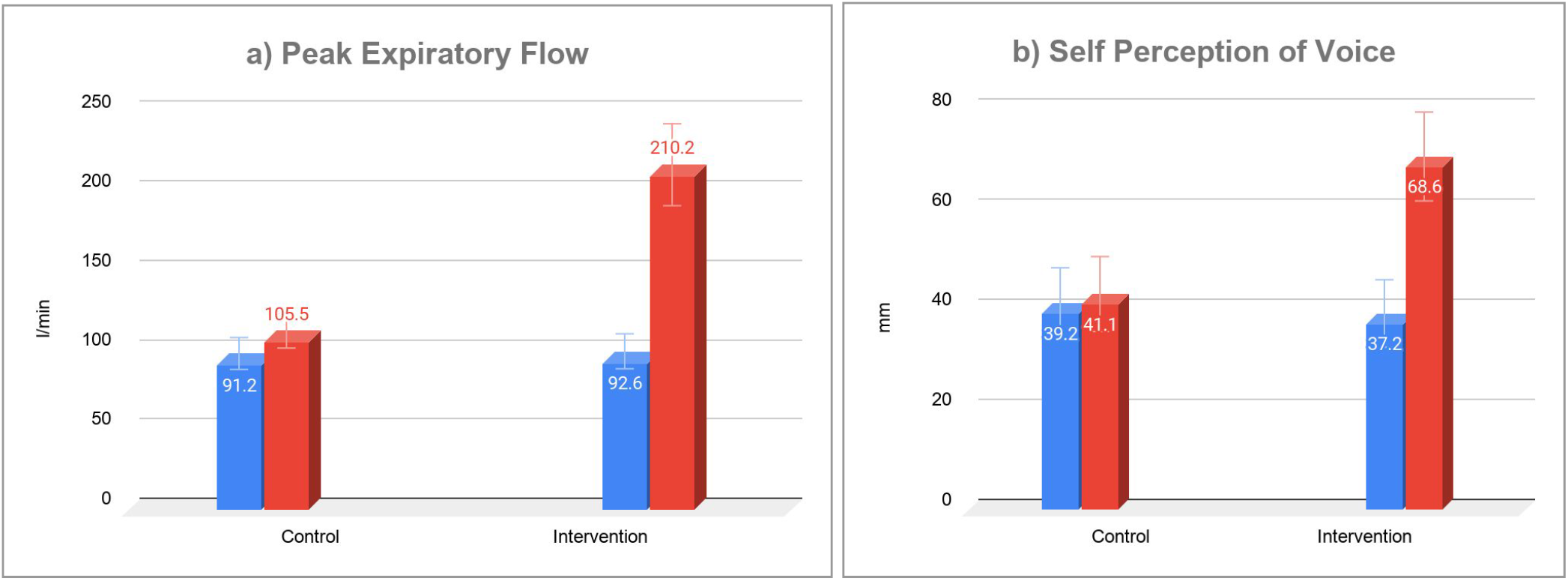
Difference in mean peak expiratory flow (a) and self perception of voice (b) before (blue bars) and after (red bars) intervention in the control (no intervention) and intervention group (cRMT intervention). Error bars indicate standard error.

### Consensus Auditory-Perceptual Evaluation of Voice (CAPE-V)

CAPE-V provides a standardized perceptual evaluation of voice [30], assessing five distinct parameters as well as perceived overall severity of the voice disorder. Table 3 shows that all parameters improved significantly in the intervention group (IG) undergoing cRMT, whereas the observed change in the control group (CG) remained below six percent in all categories (for univariate analysis of interaction between experimental condition and time see supplemental data) (Figure 2). For all the vocal parameters of the CAPE-V, the listener uses a visual analog scale (as described above), so that values from 0 to 100 are generated, with the higher the value, the more abnormal that parameter is perceived. Therefore, the individuals in the IG exhibited significant progress whereas the individuals in the CG exhibit little to no progress.

**Table 3:** Pre-test and post-test means and standard error (SEM), and percent change of parameters assessed in the Consensus Auditory-Perceptual Evaluation of Voice (CAPE-V). Overall severity as well as the five categories of CAPE-V were measured before (pre-test) and after (post-test) four weeks of cRMT (intervention group, IG) or no intervention (control group, CG). * indicates a significant post-test difference between intervention and control group. ns=not significant.

| Category |  | Pretest |  | Post test |  |  |  |
| --- | --- | --- | --- | --- | --- | --- | --- |
|  | Group | Mean | SEM | Mean | SEM | Percent Change | p-Value |
| **Overall Severity** | CG | 58.70 | 6.18 | 56.80 | 6.71 | 3.24% |  |
|  | IG | 59.00 | 6.39 | 21.70 | 5.68 | 63.22%* | < .001 |
| **Roughness** | CG | 31.80 | 10.75 | 30.50 | 11.00 | 4.09% |  |
|  | IG | 29.40 | 11.13 | 13.30 | 5.29 | 54.76% | = 0.2(ns) |
| **Breathiness** | CG | 56.80 | 6.32 | 57.20 | 6.66 | 0.70% |  |
|  | IG | 58.30 | 6.67 | 22.70 | 5.38 | 61.06%* | < .001 |
| **Strain** | CG | 41.30 | 9.32 | 39.00 | 9.92 | 5.57% |  |
|  | IG | 40.20 | 9.71 | 14.70 | 4.76 | 63.43%* | < .05 |
| **Pitch Range** | CG | 58.40 | 9.01 | 56.70 | 9.23 | 2.91% |  |
|  | IG | 58.20 | 8.50 | 30.20 | 7.15 | 48.11%* | < .05 |
| **Loudness** | CG | 59.40 | 7.32 | 57.70 | 7.50 | 2.86% |  |
|  | IG | 58.60 | 7.42 | 24.90 | 6.92 | 57.51%* | < .01 |

**Figure 2:**
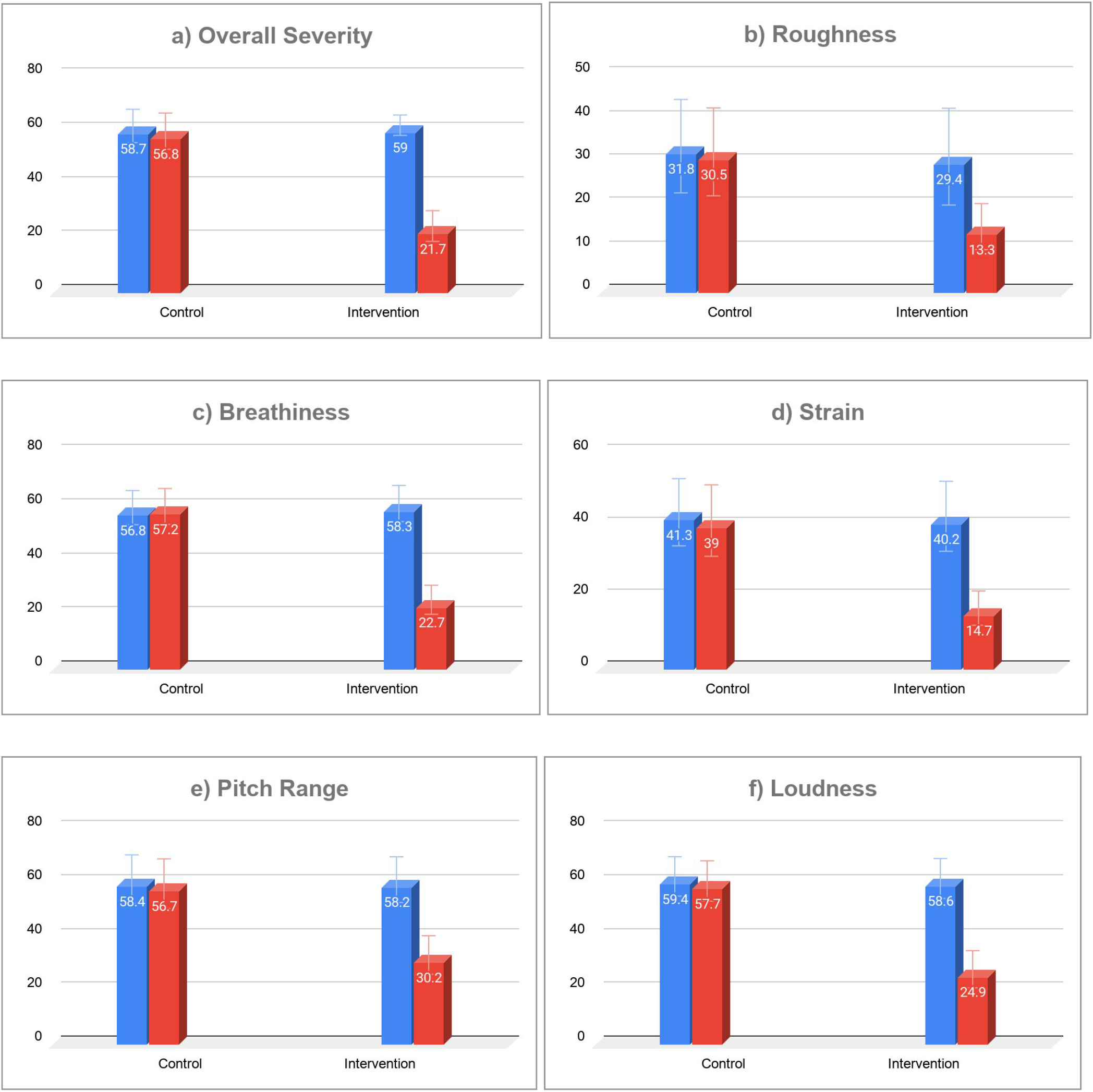
Differences in CAPE-V-assessed voice parameters. Changes are shown for overall severity (a), roughness (b), breathiness (c), strain (d), pitch range (e), and loudness (f). Blue bars show baseline data before the intervention, while red bars present post-intervention data. Patients in the control group (control) received no intervention. Subjects in the intervention group (intervention) performed 4 weeks of cRMT. Error bars indicate standard error (SEM).

### Acoustic Analysis

LSVTC software (LSVT Global, Boulder, CO) was utilized for acoustic analysis to assess the vocal intensity in dB SPL (decibels of sound pressure level) averaged over the duration of the maximum phonation time (MPT) and the vocal intensity in dB SPL in connected speech. In addition, the LSVTC software was used to assess the frequency range in Hz (Hertz) of each subject’s voice from their modal frequency to their highest frequency as well as from their modal frequency to their lowest frequency for further acoustic analysis of the effect of cRMT on voice. Table 4 shows the mean measurements of acoustic parameters before (pre-test) and after (post-test) four weeks of cRMT. Vocal intensity (dB SPL) significantly increased in the intervention group (IG) compared to the control group (CG) for both MPT (high /a/, low /a/) and connected speech (functional phrase, reading, conversation). Furthermore, the duration of MPT (duration /a/) significantly improved in the intervention vs. the control group.

**Table 4:** Pre-test and post-test means and standard error (SEM), as well as percent change. Using the LSVTC clinicians software, acoustic categories were measured before and after four weeks of cRMT (intervention group, IG) or of no intervention (control group, CG). Significant differences in post-test outcomes are indicated by *.

| LSVTC categories |  | Pretest |  | Post-test |  |  |  |
| --- | --- | --- | --- | --- | --- | --- | --- |
|  | Group | Mean | SEM | Mean | SEM | Percent Change | p-Value |
| **MPT /a/ dB SPL** | CG | 64.10 | 1.40 | 65.80 | 1.53 | 2.65% |  |
|  | IG | 63.80 | 1.35 | 79.30 | 2.40 | 24.29%* | <.001 |
| **MPT Sec Duration** | CG | 5.20 | 1.00 | 6.90 | 1.19 | 32.69% |  |
|  | IG | 4.80 | 0.93 | 15.00 | 1.56 | 212.50%* | <.001 |
| **High /a/ dB SPL** | CG | 64.30 | 1.33 | 65.40 | 1.33 | 1.71% |  |
|  | IG | 63.30 | 1.18 | 78.00 | 2.18 | 23.22% | <.001 |
| **Low /a/ dB SPL** | CG | 64.10 | 1.19 | 65.00 | 1.26 | 1.40% |  |
|  | IG | 63.70 | 1.27 | 76.40 | 2.13 | 19.94% | <.001 |
| **Functional Phrase Level dB SPL** | CG | 65.50 | 1.36 | 66.40 | 1.35 | 1.37% |  |
|  | IG | 64.50 | 1.33 | 79.50 | 2.24 | 23.26%* | <.001 |
| **Text/Word Reading Level dB SPL** | CG | 64.80 | 1.09 | 65.60 | 1.26 | 1.23% |  |
|  | IG | 63.60 | 1.39 | 77.80 | 2.20 | 22.33%* | <.001 |
| **Conversation Level dB SPL** | CG | 62.60 | 1.06 | 63.30 | 1.15 | 1.12% |  |
|  | IG | 62.50 | 1.27 | 75.60 | 2.07 | 20.96%* | <.001 |

Significant differences between groups in response to cRMT are observed in the upper pitch range (high /a/) and lower pitch range (low “Ah). Table 5 and Figure 3 also outline the increase in total pitch range, which is more pronounced in the intervention than in the control group, indicating that cRMT increased the vocal range, compared to no treatment.

**Table 5:** Pre-test and post-test means and standard error (SEM). Using the LSVTC clinicians software, acoustic categories were measured before and after four weeks of cRMT (intervention group, IG) or of no intervention (control group, CG). Changes (Δ) in semitones were calculated using [35]. Significant differences in post-test outcomes are indicated by *.

| LSVTC categories | Group | Pretest |  | Post-test |  | p-Value |
| --- | --- | --- | --- | --- | --- | --- |
| **High /a/** |  | Means | SEM | Means | SEM |  |
| **Frequency (Hz)** | CG | 159.90 | 18.66 | 161.40 | 18.34 | - |
|  | IG | 159.70 | 18.24 | 233.20 | 19.56 | - |
| **Modal Fo (Hz)** | CG | 131.90 | 16.52 | 132.40 | 16.42 | - |
|  | IG | 133.30 | 14.72 | 135.20 | 15.01 | - |
| **$\Delta$ Frequency** | CG | 28 | 2.14 | 29 | 1.92 | n.s. |
|  | IG | 26.4 | 3.52 | 98 | 4.55 | <.001 |
| **$\Delta$ Semitones** | CG | 3.3 | 0.83 | 3.4 | 0.84 | n.s. |
|  | IG | 3.0 | 0.69 | 9.6* | 1.00 | <.001 |
| **Low /a/** |  |  |  |  |  |  |
| **Frequency in Hz** | CG | 109.10 | 10.31 | 109.70 | 10.30 | - |
|  | IG | 113.20 | 9.12 | 98.40 | 6.04 | - |
| **Modal Fo (Hz)** | CG | 130.90 | 16.45 | 131.60 | 16.43 | - |
|  | IG | 133.90 | 14.87 | 135.50 | 15.05 | - |
| **$\Delta$ Frequency** | CG | 21.8 | 6.15 | 21.9 | 6.13 | n.s. |
|  | IG | 20.7 | 5.75 | 37.1 | 9.00 | n.s. |
| **$\Delta$ Semitones** | CG | 2.74 | 0.96 | 2.8 | 0.94 | n.s. |
|  | IG | 2.6 | 0.72 | 5.1 | 1.03 | n.s. |
| **Total Range** |  |  |  |  |  |  |
| **$\Delta$ Frequency** | CG | 50.8 | 8.36 | 51.7 | 8.04 | n.s. |
|  | IG | 46.5 | 9.11 | 134.8 | 13.51 | <.001 |
| **$\Delta$ Semitones** | CG | 6.2 | 1.35 | 6.3 | 1.30 | n.s. |
|  | IG | 5.5 | 1.25 | 14.6* | 1.11 | <.001 |

**Figure 3:**
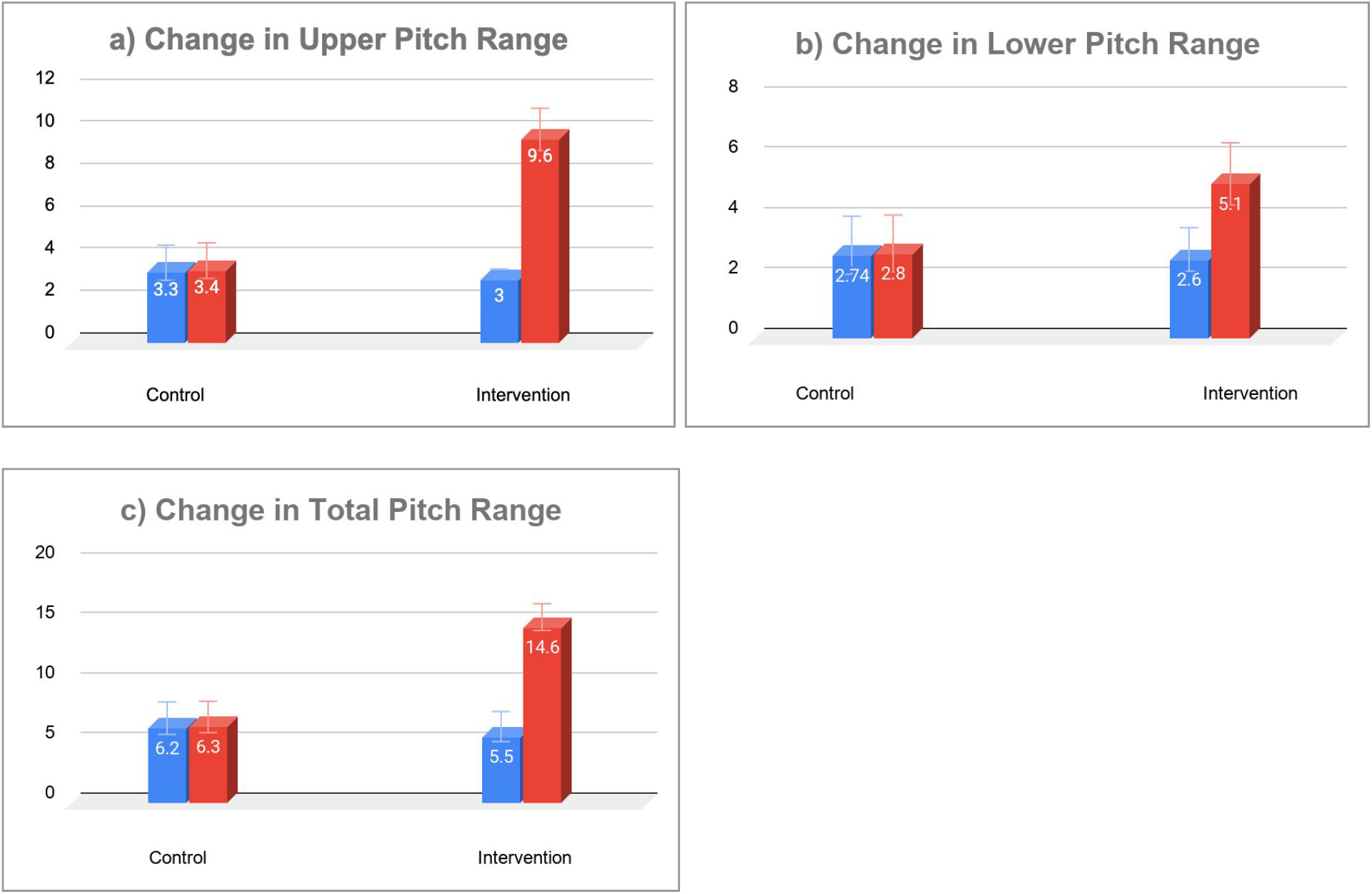
Changes in pitch range in response to cRMT (intervention) or no cRMT (control). Blue bars show differences between upper (a), lower (b) or total (c) pitch range before intervention (cRMT) or control (no treatment), while red bars show differences after four weeks or cRMT/no intervention. Error bars indicate standard error (SEM).

## Discussion

In this study, the overall data reveal cRMT is an effective therapy when compared to no treatment of neurogenic dysphonia following stroke. Significant improvements were realized with the pulmonary function test of peak flow and with the patient self-perception of vocal improvement via a Visual Analogue Scale. Improvements in peak flow observed here correspond with improved speech clarity and perception of improved voice quality, underlining the close relationship between respiratory function, phonation, and speech [13].

In addition, the detection of several notable improvements in the perceptual analysis of voice with the CAPE-V as well as some salient improvements with the acoustic analysis of voice as revealed by the LSVTC software further illustrate the usefulness of cRMT in the treatment of neurogenic dysphonia with this population. With the exception of roughness, all parameters assessed by CAPE-V improved significantly. Although cRMT does not target the intrinsic muscles of the larynx to facilitate improved glottic closure, breathiness was surprisingly noted to be reduced with cRMT. This may be due to optimization of the vibration of the true vocal folds with the resultant increased flow rate achieved with cRMT. In contrast, the CAPE-V did not observe significant improvements in roughness, as the muscles of glottic closure are not directly targeted by RMT. Improvements with strain were expected as breathing techniques, including RMT, have been shown to be effective as a stress management intervention which in turn can reduce general body neuromuscular tension, including laryngeal tension which may be increased and/or exacerbated by the stress of respiratory muscle weakness [36]. Addressing the underlying respiratory muscle weakness may facilitate a reduction of maladaptive laryngeal compensation. Increased loudness was realized as RMT targets the power source of voice with improvement of respiratory muscle strength. Unexpected improvement was noted with expansion of upward pitch range as RMT does not target the vocalis or thyroarytenoids, the primary laryngeal muscles of pitch control. However, this may be due to the increased airflow from the improved breath support resulting from cRMT rather than from improved glottic closure itself. It may have been that patients in the treatment group simply performed the upward pitch glide task with greater lung volumes, allowing them to produce phonation of longer duration and access the upper limits of their physiologic pitch range. In terms of the acoustic parameter of vocal intensity, the study revealed significant improvements across the parameters of improved vocal intensity as well as improved vocal endurance with the MPT, improved vocal intensity with glissandos from low pitch to high pitch (high /a/) as well as with glissandos from high pitch to low pitch (low /a/), and improved vocal intensity at the word, phrase, and conversation levels.

In regard to the data pertaining to frequency and pitch, examination of the acoustic parameter of frequency revealed the upward portion of the frequency and corresponding semitone ranges (From Modal Frequency and higher) exhibited significant expansion as did the total range. These findings may evidence that improvement in breath support can facilitate expansion of vocal range in this population. This contributes to expanding the pitch range of speech across the levels of word, phrase, and conversation. These marked improvements in connected speech contribute to improved intelligibility of speech by the listener. This in turn ensures the best conditions for the intended content and emotion to be transmitted successfully. The downward expansion of the frequency and corresponding semitone ranges from Modal Frequency and lower were not statistically significant. This most likely is due to the fact that a given persons’ Fundamental Frequency is often at the low end of his/her frequency range.

## Limitations

Several limitations are inherent involved in this study. The first limitation of this retrospective pilot study is the inability to employ randomization of subjects into the control and intervention groups as well as the lack of a sham therapy arm. The lack of randomization renders this study vulnerable to a skewing of the data towards positive outcomes since subject motivation may have been higher in the intervention group. Unfortunately, the level of cognitive communication competence could not be controlled for in this study. This presents the challenge of discerning whether or not the subjects in the intervention group had any better ability to process, attend, and implement the cRMT protocol used in this study. In addition, it is possible persons in the intervention group with less impaired cognitive communication competence might have been able to participate with a greater level of physical activity thus resulting in a difference in the level of engagement with the cRMT exercise compared to their counterparts in the control group. Furthermore, the average age of the intervention group was younger than the control group which may have predisposed them to have greater improvement as they may have had greater functional reserve. This is problematic as general exercise alone can facilitate improved respiratory muscle strengthening which in turn may contribute to an improved voice. Another limitation is the small sample size of this retrospective pilot study which may result in a limiting of its overall conclusiveness. Lastly, as this study did not control for smokers versus nonsmokers or for the presence and severity of laryngopharyngeal reflux (LPR), future research in this area would be enhanced by the inclusion of controlling for use of tobacco products as well as the presence and severity of acid reflux.

## Conclusions

This study analyzes the use of combined Respiratory Muscle Training (cRMT) with a resistive device simultaneously strength training both the muscles of inhalation and the muscles of exhalation. Four weeks of cRMT improved respiratory function, which in turn resulted in healthier phonation, improving the severity of dysphonia following single stroke. Therefore, this present study suggests the effectiveness, feasibility, and usefulness of inclusion of combined inspiratory muscle training and expiratory muscle training (cRMT) with a single device in the treatment of persons with neurogenic dysphonia after CVA. Further rigorous investigations evaluating the effect of cRMT on dysphonia following CVA are warranted to verify the findings of this study in larger, more diverse populations.

## Data Availability

All data available upon request

## Acknowledgements

The authors thank Sigfredo Aldarondo, MD, Tom Berlin, DHSc, MSc, RRT, and Elizabeth (Betsy) Page, MA, CCC-SLP, Sabine E. French, Ph.D., and Michael C J Quinn, PhD, for helpful comments and critical reading of the manuscript.

## Disclosures and Contributions

CG declares no conflict of interest. NB serves as independent Chief Scientist for PN Medical. RJA declares no conflict of interest

